# Increased pediatric RSV case counts following the emergence of SARS-CoV-2 are attributable to increased testing

**DOI:** 10.1101/2024.02.06.24302387

**Authors:** Brittany A. Petros, Carly E. Milliren, Pardis C. Sabeti, Al Ozonoff

## Abstract

**Background:** The incidence of respiratory syncytial virus (RSV) dropped markedly early in the COVID-19 pandemic, followed by a resurgence with heightened case counts. The “immunity debt” hypothesis proposes that the RSV-naive pediatric population increased during the period of low transmission, resulting in a subsequent increased risk of infection. However, the evidence supporting this hypothesis is limited, and no studies have comprehensively evaluated the role of changing respiratory viral testing practices in the perceived surge.

**Methods:** We conducted a multicenter, retrospective analysis of 342,530 RSV encounters and 980,546 RSV diagnostic tests occurring at 32 United States pediatric hospitals between 2013 and 2023. We used interrupted time series analysis to estimate pandemic-associated changes in RSV patient and testing volume, and to quantify changes in the proportions of patients admitted from the emergency department (ED), admitted to the intensive care unit (ICU), and receiving mechanical ventilation. We quantified the fraction of the observed shifts in case counts and in the age of diagnosed patients attributable to changes in RSV testing practices. Finally, we analyzed 524,404 influenza virus encounters and 1,768,526 influenza diagnostic tests to address the specificity of the findings to RSV.

**Findings:** RSV patient volume increased 2.4-fold (95% CI: 1.7, 3.5) in 2021-2023 relative to the pre-pandemic phase, and was accompanied by an 18.9-fold increase (95% CI: 15.0, 23.9) in RSV test volume. Over two-thirds of the apparent shifts in patient volume and in patient age were attributable to increased testing, which was concentrated among older pediatric patients. The proportions of patients with RSV requiring hospitalization, intensive care, or mechanical ventilation declined significantly across all patient age groups. These declines were not observed for patients with influenza virus.

**Interpretation:** A surge in RSV testing, rather than in viral circulation, likely underlies the increased case counts observed in 2021-2023. We identify expected consequences of increased testing, including the diagnosis of less severe cases and a shift in the patient age distribution. These findings warrant a critical assessment of the immunity debt hypothesis, while highlighting the importance of considering the testing denominator when surveillance strategies are dynamic.

**Funding:** National Institutes of Health & Howard Hughes Medical Institute

## Introduction

In 2021-2022 and 2022-2023, multiple countries experienced respiratory syncytial virus (RSV) epidemics with both altered seasonality and increased case counts relative to former RSV seasons^1–5^. The ongoing 2023-2024 epidemic in the United States is also notable for hospitalization rates that are unprecedented relative to years prior to the COVID-19 pandemic.^6^ Viral genomic analyses have shown that multiple, pre-existing RSV genotypes are circulating, suggesting that viral factors are unlikely to contribute to the observed dynamics^1,2,4^. Instead, the prevailing hypothesis remains that alterations in human behavior (e.g., social distancing and masking) during the COVID-19 pandemic reduced RSV transmission^1,5,7,8^, resulting in an increase in the pediatric population naive to RSV, i.e., the immunity debt hypothesis^9,10^.

While the immunity debt hypothesis has garnered significant attention, the evidence supporting it is limited. Multiple studies have demonstrated that pediatric RSV patients from the 2021 and 2022 seasons are older on average than in previous years^5,7,8,11–13^. However, increased age has not been mechanistically tied to an immunity debt, and was also observed among children diagnosed in 2020^14^, the year in which the debt was supposedly accumulating. While two studies have identified a decline in RSV-targeting IgG concentrations during the pandemic^15,16^, another study found no change in neutralizing antibody titers,^17^ such that the clinical or epidemiological relevance of this decline is unclear. The alternative “immunity theft” hypothesis posits that COVID-19 infections weakened our immune response to other pathogens.^18^ While one study found that children with an electronic health record (EHR) diagnosis of COVID-19 were more likely to receive an EHR diagnosis for RSV^19^, the most likely explanation for the result is that an individual’s tendency to seek healthcare is relatively constant.

The role of changing diagnostic testing practices in the large post-pandemic RSV epidemics remains to be explored. In response to COVID-19, testing capacity increased^20^, and multi-pathogen assays that simultaneously target SARS-CoV-2, influenza, and RSV were widely deployed.^21^ Indeed, one study reporting an increase in pediatric patient age noted that RSV testing increased five-fold^22^, while another stated that all individuals admitted to the hospital were tested via multi-pathogen respiratory viral tests starting in 2021^11^. Given that RSV infections with milder disease presentations – which predominantly occur in older pediatric patients – are known to escape clinical detection,^13,15^ the shift in age distribution may be a consequence of the historical underestimation of the burden of RSV.^23^ Increased respiratory viral testing can also address a major flaw of the immunity debt hypothesis, in that it appears not to have been repaid despite successive years of heightened case counts.^18^

In this study, we aim to assess the relationship between respiratory viral testing practices and RSV epidemiology, before and after the onset of the COVID-19 pandemic. We conduct a multi-center, retrospective analysis of RSV diagnostic tests and clinical encounters at 32 United States pediatric hospitals between 2013 and 2023. We quantify changes in test volume, and quantify the degree to which perceived shifts in the epidemiology of RSV can be attributed to altered testing practices.

## Methods

### Data acquisition and ethics statement

We extracted data from the Pediatric Health Information System (PHIS)^24^, which contains administrative data from a network of 48 United States tertiary care children’s hospitals spanning all 9 geographic census divisions. We isolated emergency department (ED) visits and hospitalizations associated with (i) an RSV diagnosis, (ii) an RSV diagnostic test, (iii) an influenza virus diagnosis, or (iv) an influenza diagnostic test, provided that: (i) the encounter discharge date was between July 1, 2013 and June 30, 2023, (ii) the associated hospital submitted data every month of the study period from both ED and inpatient units (32 of 48 hospitals), and (iii) the patient was under 18 years old (**Supplementary Methods**).

This study uses de-identified data and is thus considered nonhuman subjects research, exempt from requiring Institutional Review Board (IRB) approval, per the policies of Boston Children’s Hospital IRB and per an exempt determination at the Broad Institute (#NHSR-8507).

### Statistical analysis

Analyses were conducted in R v.4.1.1, with temporal data analyzed at the resolution of month of discharge date. Demographic and clinical characteristics of patients were compared between the pre-COVID-19-pandemic (“pre-pandemic”) and post-SARS-CoV-2-emergence (“post-emergence”) phases. The pre-pandemic phase was defined as ending on April 1, 2020, the date at which over 70% of states had issued stay-at-home orders.^25^ The post-emergence phase was defined as starting on the first month in which RSV patient volume exceeded that of April 2020, signifying the resumption of annual RSV epidemics. Binary variables were compared using Fisher’s exact test, continuous variables were compared using the Wilcoxon rank sum test, and categorical variables were compared using chi-square tests. Two-sample z-tests were used to test for differences in the proportions of patients within an age stratum requiring hospitalization, intensive care, or mechanical ventilation across the two phases.

We conducted interrupted time series (ITS) analysis to identify trends in encounter and testing volume, considering both linear and log-linear models with the following independent variables: (i) time, (ii) indicator variables for the intermediate period (*I*_*pand*_) and the post-emergence phase (*I*_*PE*_), (iii) variables enabling a change in slope for each phase, and (iv) harmonic terms to model seasonality (*H*_*0*_, *H*_*pand*,_ and *H*_*PE*_). We constructed models with all combinations of 0-2 harmonic terms per phase (i.e., seasonality was independently modeled for each phase using maximally one sine and one cosine term), selecting the model that minimized the transformation-adjusted Akaike Information Criterion (AIC)^26^. We also conducted ITS to estimate the effects of pandemic-related restrictions on the proportions of (i) RSV tests that were positive, (ii) ED patients admitted to the hospital, (iii) inpatients requiring intensive care, and (iv) inpatients receiving mechanical ventilation, considering linear models with up to 4 harmonic terms per phase (i.e., seasonality was independently modeled for each phase using maximally 2 sine and 2 cosine terms). For log-linear models, the equations were as follows:

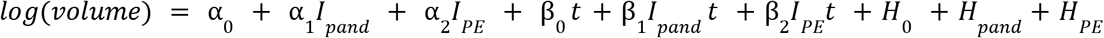

Allowing us to calculate the fold-change in the post-emergence phase, relative to the pre-pandemic phase, as follows:

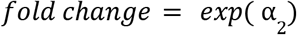

For linear models, the equations were as follows:

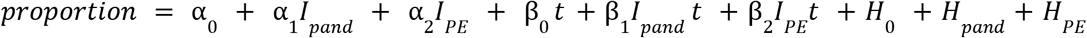

Such that the additive change in the post-emergence phase, relative to the pre-pandemic phase, was α _2_

We conducted two modified bootstrap analyses to estimate the effects of testing changes on (i) RSV patient volume and (ii) patient age. First, we estimated post-emergence test volumes under the counterfactual (i.e., no pandemic-associated disruptions) by considering linear and log-linear models of pre-pandemic test volumes as a function of time. We then subsampled post-emergence tests according to the predicted monthly test volumes, and tallied the counterfactual RSV patient volume as those who received a clinical diagnosis of RSV or those whose tests were sampled. Second, we bootstrapped the post-emergence testing data according to the age distribution (in 90-day bins) derived from the pre-pandemic testing data. Here, the counterfactual RSV patient volume consisted of those whose tests were sampled, and the counterfactual patient age distribution was analyzed.

## Results

### Increases in RSV patient volume can be attributed to changing testing paradigms

Between July 2013 and June 2023, across 32 geographically diverse pediatric hospitals, 980,546 RSV diagnostic tests were conducted and 342,530 clinical encounters associated with an RSV diagnosis occurred. We divided the study period into 2 phases by analyzing monthly patient volume: the pre-pandemic phase (July 2013 – March 2020) and the post-emergence phase (April 2021 – July 2023). No RSV epidemic occurred in the intermediate pandemic year, with a median of 46.5 monthly patients (IQR, 32–78.5) across all 32 hospitals; this period was thus excluded from further study.

We first examined temporal trends in RSV test and patient volume via interrupted time series analysis (ITS). We quantified deviations in volume that existed above and beyond the secular trends (e.g., an annual increase in RSV patient volume observed over multiple years). RSV test volume declined in the pre-pandemic phase, then demonstrated a remarkable increase by a factor of 18.9 (95% CI: 15.0, 23.9) in the post-emergence phase (**Figure 1A, Supplementary Table 1**). The bulk of the additional RSV testing was conducted via molecular tests that simultaneously detect SARS-CoV-2 and other respiratory pathogens (“SARS-CoV-2 multi-pathogen tests”; **Supplementary Figure 1**), which made up 89.8% of tests conducted in the post-emergence period. The volume of patients diagnosed with RSV increased by a factor of 2.4 (95% CI: 1.7, 3.5) in the post-emergence phase (**Figure 1B, Supplementary Table 1**). While case detection increased, test positivity dropped significantly from 14.5% (95% CI: 12.8%, 16.3%) in the pre-pandemic phase to 6.1% (95% CI: 1.9%, 10.3%) in the post-emergence phase (**Figure 1C, Supplementary Table 1**). Though the seasonality of RSV changed from the pre-pandemic to the post-emergence phase (**Figure 1D**), post-emergence patient volumes fell within the upper and lower boundaries of the 95% prediction interval derived from pre-pandemic data for 26 of 27 (96%) months.

**Figure 1.**
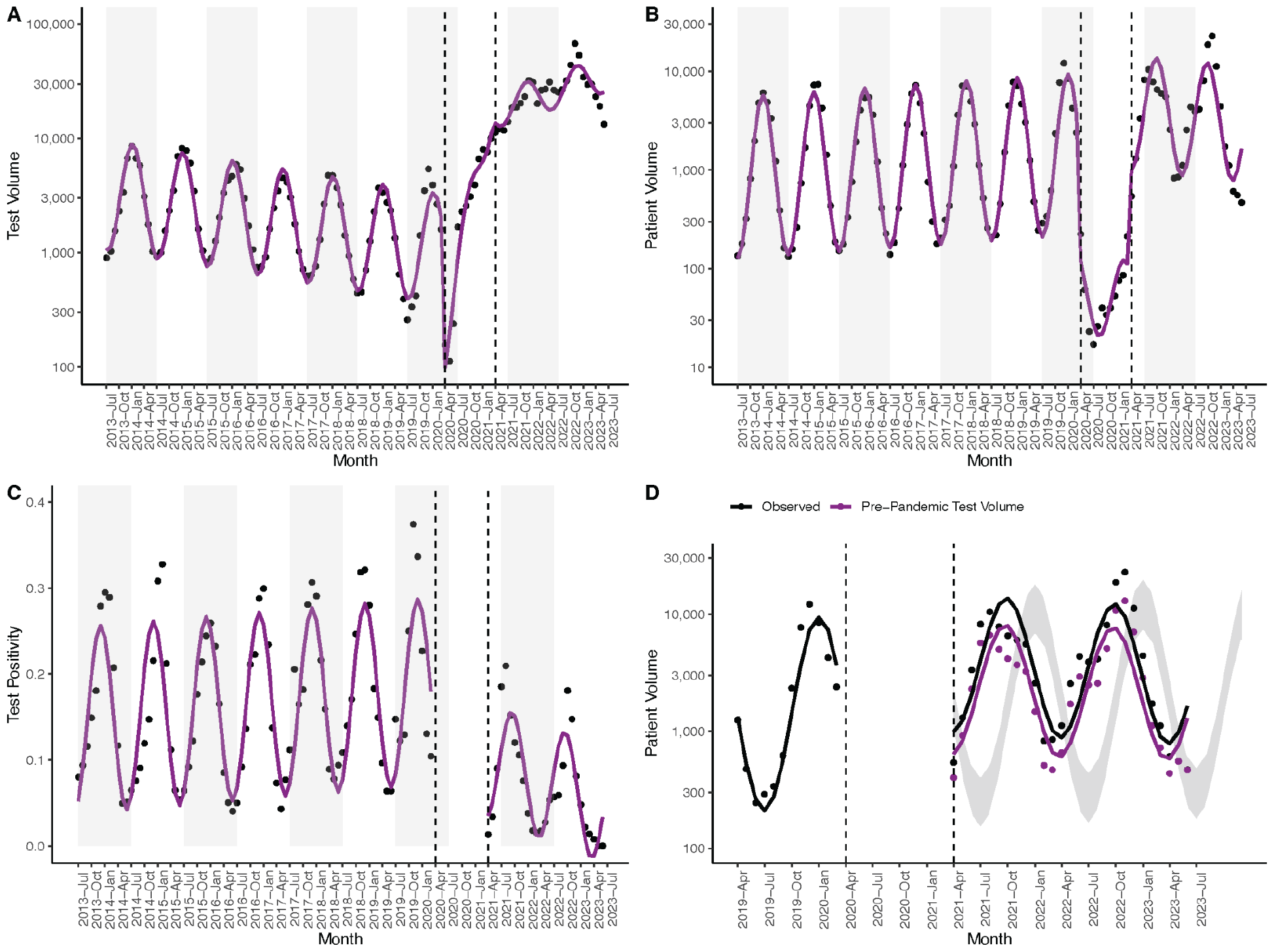
**A–C**. Monthly RSV test volume (**A**), patient volume (**B**) and test positivity (**C**). Black dots signify observed values, with model fits in magenta. Gray rectangles denote alternating years (from July – July). **D**. Monthly patient volume from April 2019 – July 2023. Observed patient volume (black) is compared to simulated patient volume under test volumes predicted via pre-pandemic testing data (magenta). Dots signify observed values, with model fits as lines. The prediction interval (gray) depicts patient volume predicted from pre-pandemic patient volumes. **A–D**. Dashed lines delineate the end of the pre-pandemic phase and the start of the post-emergence phase.

Though the SARS-CoV-2 multi-pathogen tests can also diagnose influenza virus, influenza patient volume did not increase in the post-emergence phase (**Supplementary Figure 2A, Supplementary Table 1**). While RSV test volumes increased by a factor of 18.9 (95% CI: 15.0, 23.9) in the post-emergence phase, influenza test volume increased by a factor of 3.0 (95% CI: 2.0, 4.3; **Supplementary Table 1**). In other words, influenza testing was more robust than RSV testing prior to the emergence of SARS-CoV-2, with the introduction of SARS-CoV-2 multi-pathogen tests resulting in a smaller increase in influenza test volume (**Supplementary Figure 2B)**.

To assess the impact of increased testing on increased RSV patient volume, we simulated a counterfactual scenario in which post-emergence test volume was consistent with pre-pandemic test volume (**Methods**). Under this scenario, RSV patient volume increased by a factor of 1.5 (95% CI: 1.0, 2.0) in the post-emergence phase (**Figure 1D, Supplementary Table 1**), such that 68.3% of the observed increase in patient volume can be attributed to increased testing. In summary, both RSV testing and patient volume increased in the post-emergence phase, with changes in volume largely attributable to changes to testing.

### Increased testing of older children is associated with increased patient age

We next compared the demographic and clinical characteristics of patients tested for and diagnosed with RSV in the pre-pandemic and post-emergence phases. Patients tested for RSV differed across the phases among multiple demographic axes (**Table 1**), the most striking of which was age, which increased from a median of 8.6 months to 35.1 months (p < 0.001, **Table 1, Figure 2A, Supplementary Figure 3**). Patients tested for RSV were also significantly less likely to have a complex chronic condition^27^ (CCC) in the post-emergence phase, defined as a medical condition expected to last at least 12 months, requiring specialty care and periods of hospitalization at tertiary care centers^28^ (15.8% vs. 18.5%, p < 0.001; **Table 1**). A larger proportion of RSV tests were conducted on older and healthier children in the post-emergence phase, and test positivity dropped significantly across all age groups (p < 0.001; **Table 1**).

**Figure 2.**
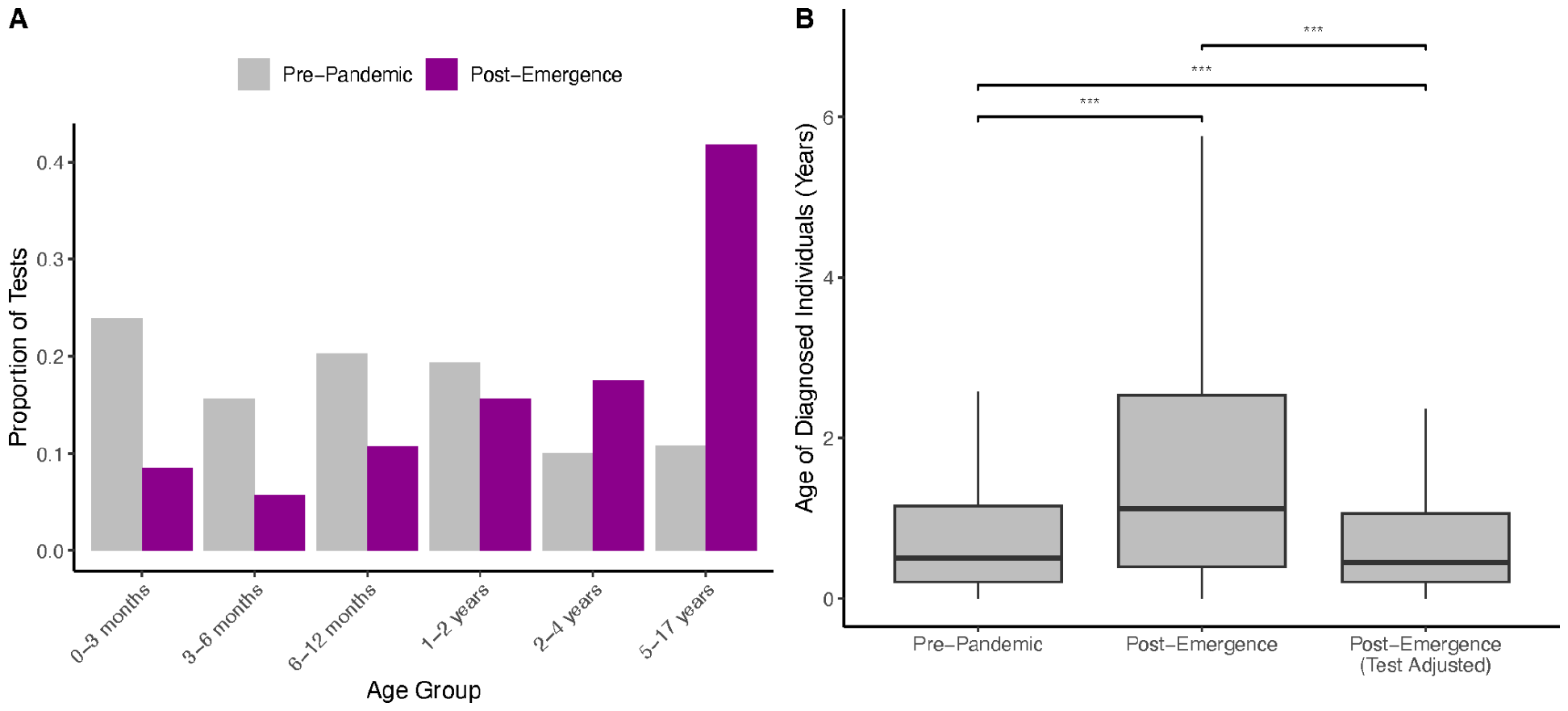
**A**. Proportion of RSV tests conducted on each age group in the pre-pandemic (gray) vs. post-emergence (magenta) phase. **B**. Age, in years, of patients tested for and diagnosed with RSV in the pre-pandemic and post-emergence phases. Because tests in the post-emergence phase were conducted on older patients, on average, than in the pre-pandemic phase, the post-emergence testing data was bootstrapped according to the age distribution of the pre-pandemic testing data. The resulting patient age distribution is reported as “Post-Emergence (Test Adjusted).” Boxplots display the first, second, and third quartiles, with whiskers extending to the minimum of 1.5 times the interquartile range and the most extreme data point in either direction. ***, p < 0.001 via Wilcoxon rank sum test.

**Table 1.** Demographic characteristics of patients tested for respiratory syncytial virus. Medians (with interquartile ranges) are used to summarize distributions of continuous variables. ED, emergency department. IP, inpatient. NA, not applicable. PCR, polymerase chain reaction.

|  | **Pre-pandemic phase**<br>Jul 2013 – Mar 2020<br>**N = 210,424 tests** | **Post-emergence phase**<br>Apr 2021 – Jun 2023<br>**N = 723,460 tests** | **P-value** |
| --- | --- | --- | --- |
| Age (months) | 8.57 (3.12 – 20.01) | 35.05 (11.99 – 88.21) | < 0.001 |
| Age Category (%):<br>• 0-90 days<br>• 3-6 months<br>• 6-12 months<br>• 1-2 years<br>• 2-4 years<br>• 5-17 years | • 23.91%<br>• 15.63%<br>• 20.29%<br>• 19.38%<br>• 9.99%<br>• 10.81% | • 8.50%<br>• 5.73%<br>• 10.77%<br>• 15.63%<br>• 17.57%<br>• 41.80% | < 0.001 |
| Sex (% male) | 55.84% | 54.19% | < 0.001 |
| Race (%)<br>• White<br>• Black<br>• Asian<br>• Pacific Islander<br>• American Indian<br>• Other<br>• Multiple<br>• Unknown | • 43.94%<br>• 24.00%<br>• 2.31%<br>• 0.35%<br>• 0.23%<br>• 9.91%<br>• 1.15%<br>• 18.11% | • 48.89%<br>• 23.07%<br>• 4.33%<br>• 0.45%<br>• 0.35%<br>• 13.73%<br>• 3.22%<br>• 5.96% | < 0.001 |
| Ethnicity (%)<br>• Hispanic or Latino<br>• Not Hispanic or Latino<br>• Unknown | • 31.59%<br>• 62.80%<br>• 5.61% | • 30.67%<br>• 63.81%<br>• 5.51% | < 0.001 |
| Complex Chronic Condition by Age Group (%)<br>• 0-90 days<br>• 3-6 months<br>• 6-12 months<br>• 1-2 years<br>• 2-4 years<br>• 5+ years | 18.45%<br>• 13.77%<br>• 10.55%<br>• 11.39%<br>• 14.64%<br>• 27.68%<br>• 51.75% | 15.84%<br>• 14.58%<br>• 17.38%<br>• 11.62%<br>• 10.02%<br>• 8.50%<br>• 2.23% | < 0.001<br>• < 0.001<br>• < 0.001<br>• < 0.001<br>• < 0.001<br>• < 0.001<br>• < 0.001 |
| Premature or Neonatal Complex Chronic Condition (%) | 2.71% | 1.64% | < 0.001 |
| Patient Type<br>• ED (Discharge)<br>• ED & IP (ED Admit)<br>• IP (Direct Admit) | • 56.60%<br>• 34.73%<br>• 8.67% | • 66.34%<br>• 27.00%<br>• 6.66% | < 0.001 |
| Diagnostic Test (%)<br>• Culture<br>• Antigen<br>• PCR<br>• DNA Probe<br>• SARS-CoV-2 Multi-Pathogen<br>• Other/Unspecified Test<br>• Multiple Tests | • 3.05%<br>• 83.39%<br>• 5.26%<br>• 4.77%<br>• 0%<br>• 0.01%<br>• 3.52% | • 0.26%<br>• 7.52%<br>• 10.78%<br>• 0.14%<br>• 80.78%<br>• < 0.01%<br>• 0.52% | < 0.001 |
| Test Positivity by Diagnostic (%)<br>• Culture<br>• Antigen<br>• PCR<br>• DNA Probe<br>• SARS-CoV-2 Multi-Pathogen | 21.50%<br>• 9.64%<br>• 23.03%<br>• 18.16%<br>• 7.68%<br>• NA | 7.48%<br>• 9.24%<br>• 12.02%<br>• 10.97%<br>• 6.20%<br>• 6.59% | < 0.001<br>• 0.615<br>• < 0.001<br>• < 0.001<br>• 0.090<br>• NA |
| Test Positivity by Age Group (%)<br>• 0-90 days<br>• 3-6 months<br>• 6-12 months<br>• 1-2 years<br>• 2-4 years<br>• 5+ years | • 26.30%<br>• 27.47%<br>• 22.62%<br>• 20.35%<br>• 18.03%<br>• 5.46% | • 14.58%<br>• 17.38%<br>• 11.62%<br>• 10.02%<br>• 8.50%<br>• 2.23% | • < 0.001<br>• < 0.001<br>• < 0.001<br>• < 0.001<br>• < 0.001<br>• < 0.001 |

Among individuals diagnosed with RSV, patient age also increased, from a median of 6.9 months to 11.9 months (p < 0.001, **Table 2, Figure 2B**). Patients diagnosed with RSV were significantly less likely to have a CCC in the post-emergence phase (11.7% vs. 16.8%, p < 0.001; **Table 2**). Moreover, the apparent clinical severity of RSV cases declined in the post-emergence phase, with patients less likely to be admitted from the ED (38.7% vs. 50.6%); to require intensive care (27.2% vs. 32.3%), mechanical ventilation (7.4% vs. 11.9%), or extracorporeal membrane oxygenation (0.15% vs. 0.22%); or to expire in the hospital (0.16% vs. 0.24%; **Table 2**).

**Table 2.**
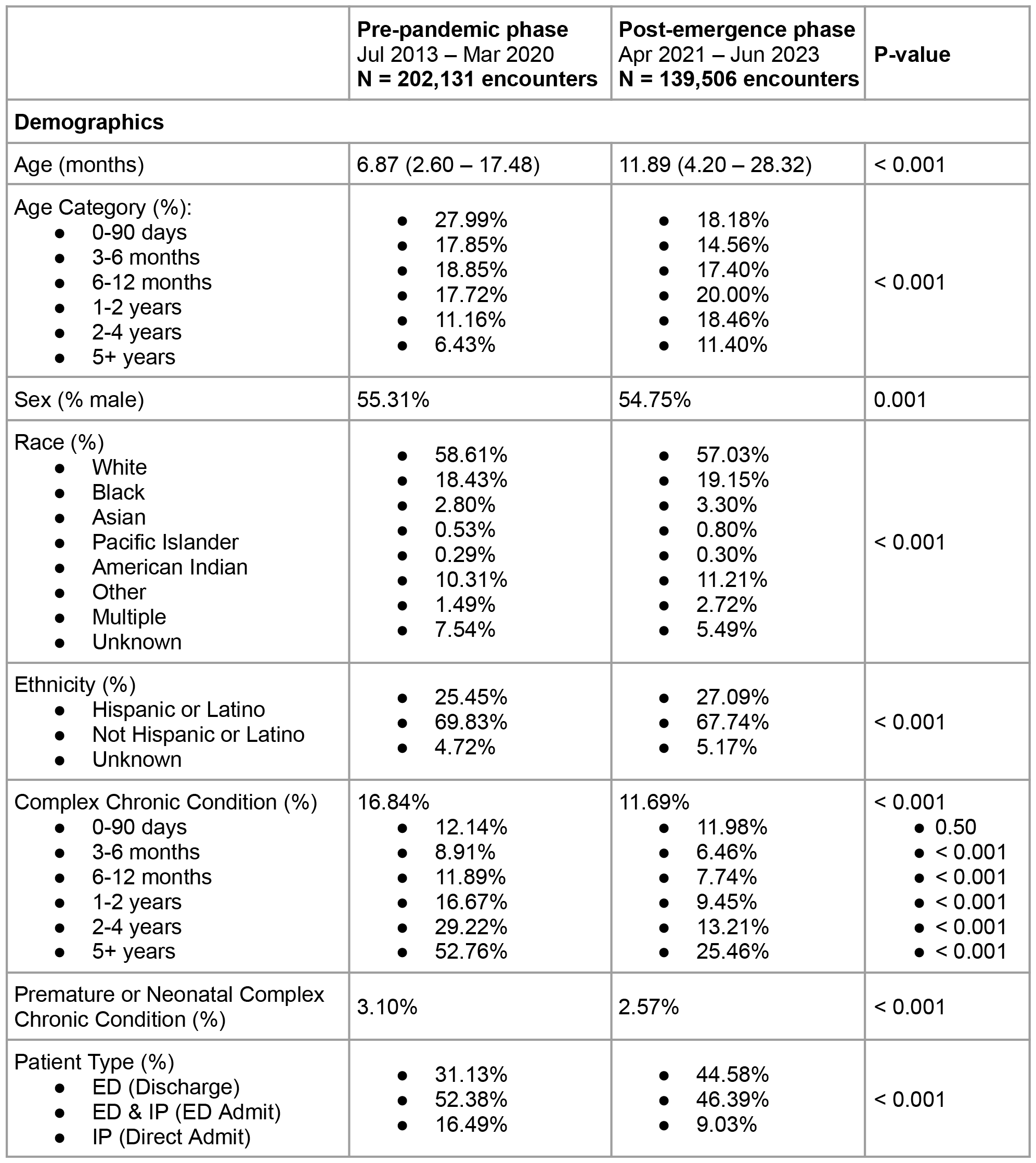
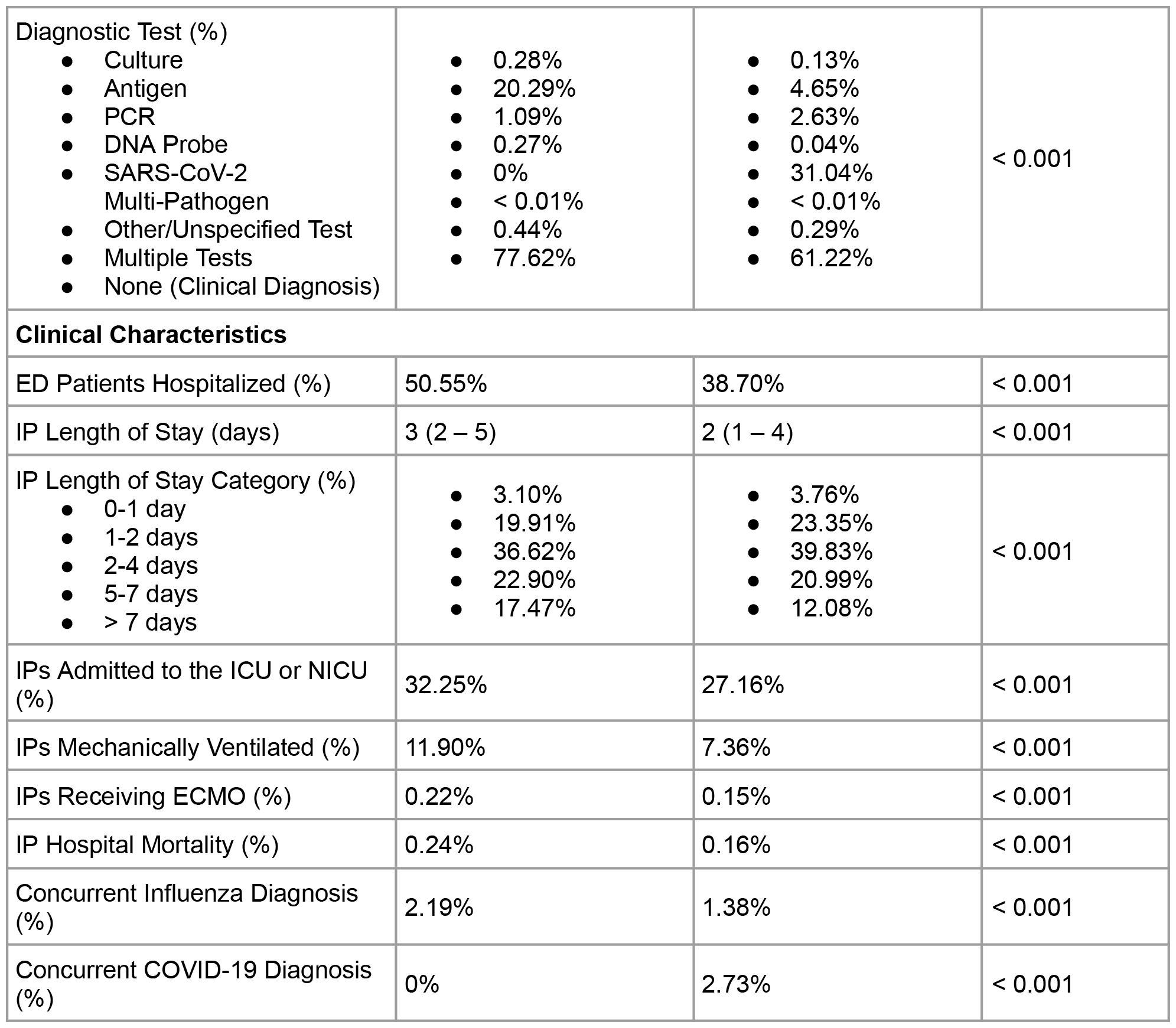
Demographic and clinical characteristics of patients diagnosed with respiratory syncytial virus. Medians (with interquartile ranges) are used to summarize distributions of continuous variables. ECMO, extracorporeal membrane oxygenation. ED, emergency department. IP, inpatient. ICU, intensive care unit. NICU, neonatal intensive care unit. PCR, polymerase chain reaction.

Among pediatric patients, the youngest patients^29–31^ are at the greatest risk for severe disease; therefore, true shifts in patient age could meaningfully impact RSV morbidity. However, we hypothesized that the apparent shifts were attributable to shifts in the age of those tested (**Figure 2A, Supplementary Figure 3**). We bootstrapped post-emergence tests according to the age distribution of the pre-pandemic testing data (**Methods**) to assess the impact of increased testing of older children on the age of children diagnosed with RSV. Under this scenario, in which the age of patients tested for RSV was held constant across phases (**Supplementary Figure 3**) and a diagnostic test was a prerequisite to RSV diagnosis, the age of patients diagnosed with RSV actually declined, from a median of 6.0 months (IQR: 2.5 – 13.9) in the pre-pandemic phase to 5.5 months (IQR, 2.5 – 12.8) in the post-emergence phase (p < 0.001; **Figure 2B**). Thus, the apparent increase in median patient age can be ascribed to changes in the age of patients tested.

### The apparent declines in clinical severity are specific to RSV

Declining measures of clinical severity were observed among RSV cases (**Table 2**), and may reflect increased detection of cases with milder disease presentations or decreased availability of healthcare resources in the context of SARS-CoV-2. We therefore examined temporal trends in clinical severity for patients with RSV and for patients with influenza virus. The fraction of patients admitted from the ED dropped from 52.6% (95% CI: 50.6%, 54.6%) in the pre-pandemic phase to 37.7% (95% CI: 32.6%, 42.9%) in the post-emergence phase for RSV (**Figure 3A, Supplementary Table 2**). In contrast, the proportion of patients diagnosed with influenza virus that were admitted from the ED increased in the post-emergence phase (**Supplementary Figure 4A, Supplementary Table 2**). The proportion of inpatients admitted to the ICU also declined among those diagnosed with RSV, from 26.3% (95% CI: 24.5%, 28.2%) to 10.4% (95% CI: 6.1%, 14.7%) (**Figure 3B, Supplementary Table 2**), but remained stable for patients with influenza (**Supplementary Figure 4B, Supplementary Table 2**). Moreover, the fraction of patients with RSV receiving mechanical ventilation declined by 10.3% (95% CI: -14.1%, -6.5%) (**Figure 3C, Supplementary Table 2**), though there was no change among those with influenza (**Supplementary Figure 4C, Supplementary Table 2**). In summary, patients diagnosed with RSV, but not influenza virus, displayed milder disease presentations in the post-emergence phase.

**Figure 3.**
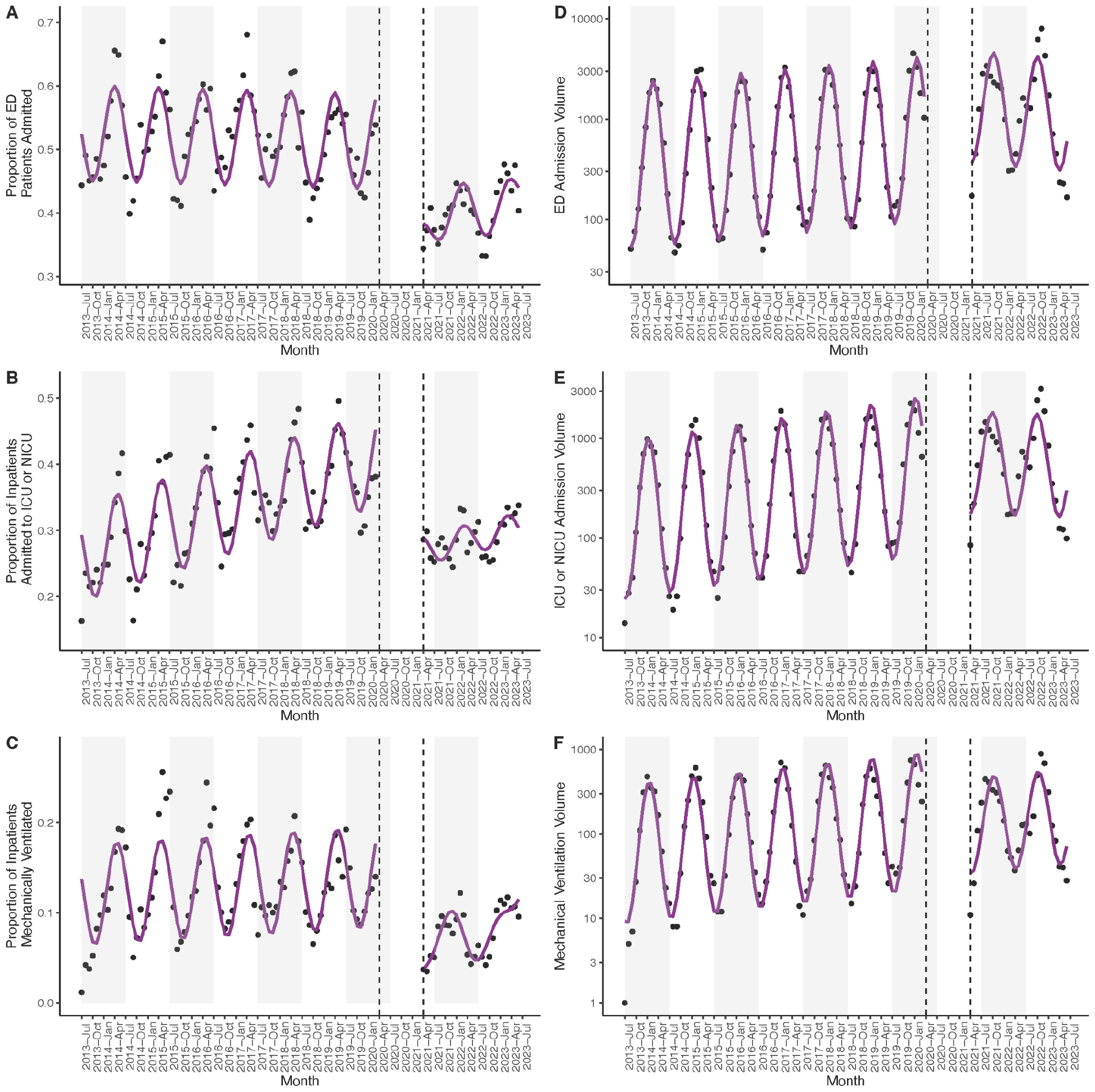
**AD**. Proportion (**A**) and volume (**D**) of patients with a diagnosis of respiratory syncytial virus (RSV) admitted from the emergency department (ED). **BE**. Proportion (**B**) and volume (**E**) of inpatients with a diagnosis of RSV admitted to the intensive care unit (ICU) or neonatal intensive care unit (NICU). **CF**. Proportion (**C**) and volume (**F**) of inpatients with a diagnosis of RSV mechanically ventilated. **A-F**. Black dots signify observed values, with model fits in magenta. Gray rectangles denote alternating years (from July – July). Dashed lines delineate the end of the pre-pandemic phase and the start of the post-emergence phase. Data in the interim period is not shown.

### The volume of severe RSV cases remains stable

We also analyzed the volume of RSV patients demonstrating severe disease, as elevated patient volumes have the potential to overwhelm healthcare systems. Both overall RSV patient volume and the volume of patients hospitalized from the ED increased in the post-emergence phase, by 2.4-fold (95% CI: 1.7, 3.5; **Figure 1B**) and 1.9-fold (95% CI: 1.3, 2.9; **Figure 3D, Supplementary Table 1**), respectively. However, the volume of patients with RSV requiring intensive care (95% CI: 0.80, 1.7) or mechanical ventilation (95% CI: 0.49, 1.2) remained stable in the post-emergence phase relative to the pre-pandemic phase (**Figure 3EF, Supplementary Table 1**). While significantly more RSV cases were detected in the post-emergence phase, there is no evidence for an increase in the number of severe cases.

### Apparent declines in RSV clinical severity are present across patient age strata

Given known relationships between RSV severity and pediatric patient age^29–31^, it is possible the declining measures of clinical severity could be attributed solely to the observed increase in patient age. However, we observed significant declines in the proportions of patients admitted from the ED, admitted to the ICU or NICU, and receiving mechanical ventilation within every age group (**Figure 4**). Among patients with RSV evaluated in the ED, the percent change in the admission proportion declined monotonically with patient age, from 7.61% (0-3 months) to 46.08% (5-17 years) (**Figure 4A**; **Supplementary Table 3**). Similarly, the percent change in the proportion of inpatients admitted to intensive care decreased monotonically with patient age, from 2.87% (0-3 months) to 25.23% (5-17 years) (**Figure 4B**; **Supplementary Table 3**), and the greatest declines in the proportion of inpatients mechanically ventilated occurred among children over 6 months of age (**Figure 4C**; **Supplementary Table 3**). Though present across age groups, reductions in clinical severity were most pronounced among older patients, reflecting the pre-pandemic practice of primarily testing older children that exhibited severe disease.

**Figure 4.**
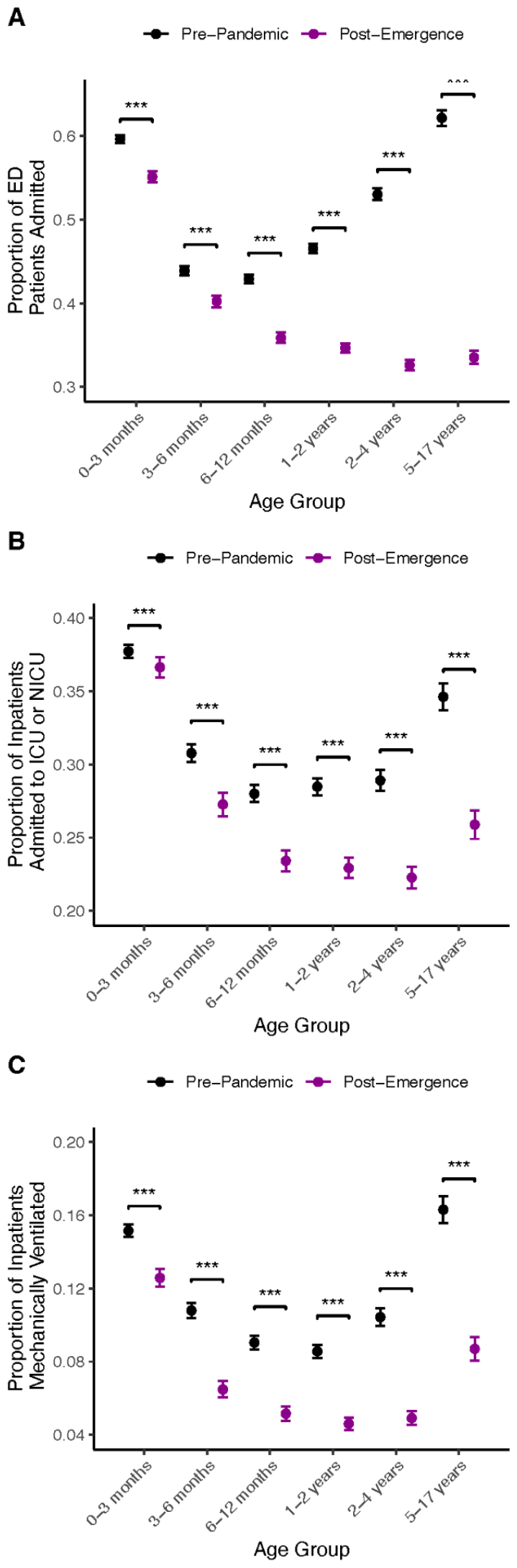
**ABC**. Proportions of emergency department (ED) patients admitted to the hospital (**A**), inpatients admitted to the intensive care unit (ICU) or neonatal ICU (NICU) (**B**), or inpatients receiving mechanical ventilation (**C**) for patients with a diagnosis of respiratory syncytial virus (RSV), by age group and phase. *, p < 0.05. **, p < 0.01. ***, p < 0.001.

## Discussion

Using administrative data from 32 pediatric hospitals in the United States, we quantified changes in the epidemiology of RSV over the last decade. While RSV patient volume doubled after the lifting of COVID-19 pandemic mitigation measures, we found that RSV test volume increased 18-fold. We identified an increase in patient age that can be largely explained by the increased testing of older children. We also documented declines in measures of clinical acuity, including the proportions of patients requiring hospitalization, intensive care, and mechanical ventilation, across all patient age groups.

Increased patient volume and an apparent decline in clinical severity are expected consequences of large increases in RSV testing. Patients with the most severe clinical courses rarely escape detection, with increased testing predominantly capturing those with more mild illness and decreasing the apparent severity of infection regardless of age (**Figure 5**). Because the risk of severe RSV declines with increased pediatric patient age^29–31^, the detection of more mild cases shifts the average patient age upwards. We found that the proportion of clinically severe cases was remarkably high among children 5-17 years old in the pre-pandemic phase (**Figure 4**), reflecting underascertainment of milder RSV infections prior to the emergence of COVID-19.^23^ Moreover, we identify increased detection of RSV in between annual epidemics in the post-emergence phase (**Figure 1B**), likely due to heightened RSV testing regardless of the season (**Figure 1A**).

**Figure 5.**
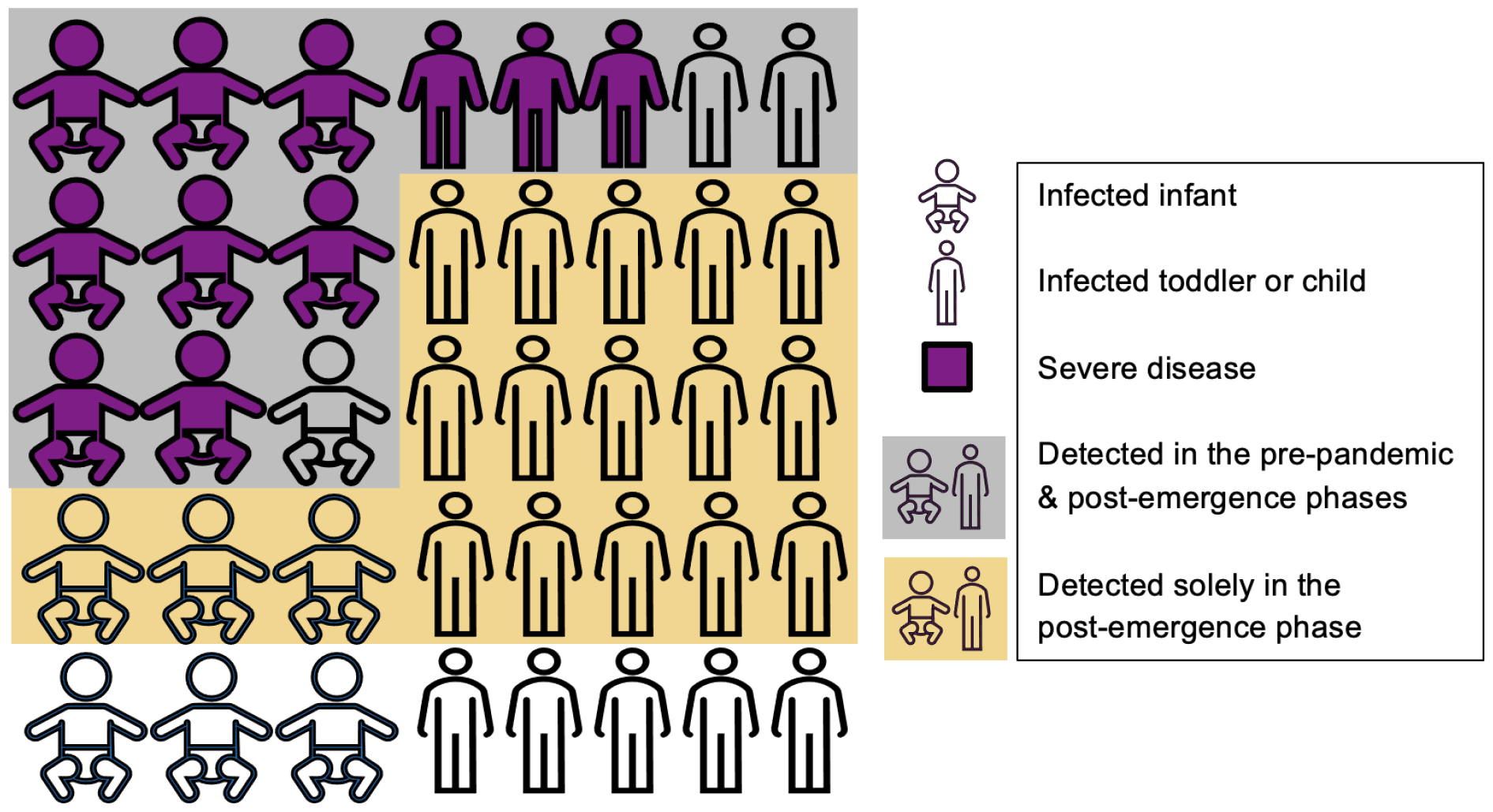
Schematic depiction of the manuscript’s conclusions. The most severe cases of respiratory syncytial virus (RSV), depicted in magenta, were detected in the pre-pandemic phase and continued to be detected in the post-emergence phase. Thus, there was no increase in the volume of patients requiring intensive care or mechanical ventilation. However, additional testing in the post-emergence phase resulted in the additional detection of primarily mild or moderate RSV cases, leading to greater patient volume. The average age of diagnosed individuals increased, and the fraction of patients experiencing clinically severe outcomes decreased, as infections in older children, who are at lower risk for severe disease, were less likely to be detected in the pre-pandemic phase.

We considered multiple other possible explanations for our results. While it remains possible that RSV circulation simultaneously increased due to decreased population-level immunity, this hypothesis is difficult to reconcile with the finding that the volume of patients requiring intensive care or mechanical ventilation was unchanged. Indeed, the clinical significance of the immunity debt paradigm remains unknown, and may be difficult to quantify given the concurrent surge in RSV testing. The observed decline in RSV clinical severity is also unlikely to be the result of hospital capacity limitations, given that no equivalent decline was noted for patients with influenza virus. Additionally, while the increase in patient age may also be impacted by a pandemic-associated increase in the tendency to seek healthcare for respiratory symptoms, testing alone can explain the increase.

There are limitations to our work. Our study was observational, and the associations that we identify may or may not be causal. PHIS contains data on tertiary-care pediatric hospitals, which may not generalize to all hospitals caring for pediatric patients with RSV. We defined our patient cohorts using ICD-9 and ICD-10 codes present in billing data, which may underestimate or overestimate the true patient volumes. Furthermore, we cannot discern ED visits and admissions specifically for RSV-related symptoms, and it is possible that some of the identified cases were incidental.

Nevertheless, our work has an important takeaway that seems to have been unaddressed in recent literature regarding RSV activity in the United States. Endemic pathogen surveillance is complex, and case surges are often contemporaneous with testing surges^32^. It thus becomes essential that we consider the shifting testing denominator when comparing metrics such as case counts to those collected in the past. As testing becomes more widespread – such that an increased fraction of the true infections is detected – we move closer to the ground truth in terms of case severity and fatality rates. In the meantime, approaches such as wastewater-based epidemiology^33,34^ can mitigate the biases induced by changes in testing, and will be important data sources for improved surveillance of RSV and other respiratory pathogens.

## Data Availability

Data analyzed in this manuscript were downloaded from the Pediatric Health Information System (PHIS) database (https://www.childrenshospitals.org/content/topics/data-insights), which prohibits data sharing outside of its member hospitals.

## Acknowledgements

This work was supported by the National Institute of General Medical Sciences (T32GM00773 and T32GM144273, to B.A.P.), the Howard Hughes Medical Institute Investigator Program (to P.C.S.) and the National Institute of Allergy and Infectious Diseases (U19AI110818, to P.C.S.). The content is solely the responsibility of the authors and does not necessarily represent the official views of the National Institute of General Medical Sciences or the National Institutes of Health. ChatGPT 3.5 was used for coding assistance, though all code underwent manual verification, annotation, and rigorous testing prior to implementation. Roby Bhattacharyya, Bronwyn MacInnis, and Gage Moreno provided thoughtful feedback on this work.

