## Supplementary Information for "Increased pediatric RSV case counts following the emergence of SARS-CoV-2 are attributable to increased testing"

### Supplementary Methods

#### Data acquisition

Emergency department (ED) visits and hospitalizations associated with an RSV diagnosis (i.e., International Classification of Diseases (ICD) 9th Revision codes 079.6, 466.11, and 480.1 or ICD 10th Revision codes B97.4, J21.0, J20.5, and J12.1) were extracted from the Pediatric Health Information System (PHIS) on September 5, 2023. ED visits and hospitalizations associated with an influenza diagnosis (ICD-9 codes 487.0, 487.1, 487.8, 488.01, 488.02, 488.09, 488.11, 488.12, 488.19, 488.81, 488.82, or 488.89 and ICD-10 codes of J09.X1, J09.X2, J09.X3, J09.X9, J10.00, J10.01, J10.08, J10.1, J10.2, J10.81, J10.82, J10.83, J10.89, J11.00, J11.08, J11.1, J11.2, J11.81, J11.82, J11.83, or J11.89) were extracted on October 24, 2023. Encounters associated with an RSV test were extracted on November 9, 2023, using the following Clinical Transaction Classification (CTC) codes: 364240.1.1.00, 364240.3.1.03, 364242.1.1.14, 364242.1.1.26, 364242.1.1.27, 364242.1.1.35, 364242.3.1.14, 364242.3.1.26, 364243.1.1.14, 364243.1.1.27, 364243.1.1.38, 364243.3.1.14, 364249, 382317.1.1.00, 382317.3.1.00, 382318.1.1.00, 382318.3.1.00, 382319.1.1.00, 382319.3.1.00, 382320.1.1.00, 382320.3.1.00, or 382350.3.2.14. These codes include RSV antigen tests, viral culture, molecular assays, and multipanel respiratory virus tests. Encounters associated with an influenza test were extracted on December 13, 2023, using the following CTC codes: 364150.1.1.00, 364150.3.1.00, 364152.1.1.26, 364152.1.1.27, 364152.3.1.26, 364152.3.1.27, 364153.1.1.27, 364153.3.1.27, 364159.1.1.00, 364159.3.1.00, 382237.1.1.00, 382237.3.1.00, 382238.1.1.00, 382238.3.1.00, 382239.1.1.00, 382239.3.1.00, 382240.1.1.00, 382240.3.1.00, or 382350.3.2.14. These codes include influenza antigen tests, molecular assays, and multipanel respiratory virus tests.

#### Data cleaning

Diagnostic codes were used to annotate RSV encounters with active influenza (ICD codes above) or COVID-19 (ICD-10 code U07.1) co-infections.

Same-day readmissions occurred in 4,840 individuals, and most commonly represented within-hospital unit transfers (e.g., a transfer from the ED to an inpatient service). These readmissions were grouped into a single encounter with the admission dates and length of stay adjusted to encompass the entire admission. If a patient was admitted to the ED, discharged, and admitted as an inpatient on the same day, the patient was considered to be admitted from the ED.

ED dispositions were grouped into the following categories: admitted as an inpatient to the hospital and other (of which 98.6% were “discharge to home or self care”, 0.4% were unknown, and 1.0% encompassed 20 distinct other dispositions). One encounter with a negative admit age was dropped. Admit ages in days were converted to admit ages in months by dividing by 30.44. Encounters occurring in the observation unit were grouped with inpatient encounters. Individuals with more than one race indicated were categorized as “Multiple.” Encounters with missing race or ethnicity information were removed when conducting demographic comparisons (i.e., missing data was assumed to be missing at random).

### Supplementary Tables

#### Supplementary Table 1

**Supplementary Table 1.** Parameter estimates during the pre-pandemic and post-emergence phases. For the log-linear models of volume, intercepts are multiplicative constants, while slopes are expressed as annual percent changes. For the linear model of test positivity, intercepts and annual slopes are additive. ICU, intensive care unit. NICU, neonatal intensive care unit. *, p < 0.05. **, p < 0.01. ***, p < 0.001.

| **Parameter** | **Estimate (95% Confidence Interval)** |
|---|---|
| **RSV Patient Volume** | |
| Pre-pandemic slope | 8.67 (4.15, 13.38)*** |
| Post-emergence intercept change | 2.42 (1.68, 3.47)*** |
| Post-emergence slope change | -18.09 (-34.89, 3.04) |
| **RSV Test Volume** | |
| Pre-pandemic slope | -14.99 (-17.29 , -12.63)*** |
| Post-emergence intercept change | 18.95 (15.00, 23.94)*** |
| Post-emergence slope change | 63.16 (40.66, 89.25)*** |
| **Test Positivity** | |
| Pre-pandemic intercept | 0.145 (0.128, 0.163)*** |
| Pre-pandemic slope | 0.005 (0.001, 0.010)* |
| Post-emergence intercept change | -0.084 (-0.122, -0.046)*** |
| Post-emergence slope change | -0.029 (-0.053, -0.004)* |
| **RSV Patient Volume under Pre-Pandemic Testing Volume** | |
| Pre-pandemic slope | 8.67 (4.35, 13.17)*** |
| Post-emergence intercept change | 1.45 (1.03, 2.05)* |
| Post-emergence slope change | -12.20 (-29.49, 9.34) |
| **Influenza Patient Volume** | |
| Pre-pandemic slope | 14.33 (3.91, 25.80)** |
| Post-emergence intercept change | 0.09 (0.04, 0.21)*** |
| Post-emergence slope change | 323.6 (152.7, 710.1)*** |

| | |
|---|---|
| **Influenza Test Volume** | |
| Pre-pandemic slope | 7.08 (2.52, 11.86)** |
| Post-emergence intercept change | 2.96 (2.04, 4.27)*** |
| Post-emergence slope change | 46.64 (16.37, 84.79)** |
| **RSV Emergency Department Admission Volume** | |
| Pre-pandemic slope | 9.46 (4.48, 14.67)*** |
| Post-emergence intercept change | 1.92 (1.29, 2.85)** |
| Post-emergence slope change | -15.93 (-34.63, 8.11) |
| **RSV ICU or NICU Admission Volume** | |
| Pre-pandemic slope | 16.71 (11.57, 22.09)*** |
| Post-emergence intercept change | 1.17 (0.80, 1.72) |
| Post-emergence slope change | -17.69 (-35.49, 5.02) |
| **RSV Mechanical Ventilation Volume** | |
| Pre-pandemic slope | 13.71 (7.82, 19.93)*** |
| Post-emergence intercept change | 0.77 (0.49, 1.21) |
| Post-emergence slope change | -1.01 (-25.75, 31.98) |

Supplementary Table 2

**Supplementary Table 2.** Parameter estimates during the pre-pandemic and post-emergence phases. For the linear models of proportions, intercepts and annual slopes are additive. ICU, intensive care unit. NICU, neonatal intensive care unit. *, p < 0.05. **, p < 0.01. ***, p < 0.001.

| **Parameter** | **Respiratory Syncytial Virus (RSV)** | **Influenza Virus** |
|---|---|---|
| **Proportion of Emergency Department Patients Admitted** | | |
| Pre-pandemic intercept | 0.526 (0.506, 0.546)*** | 0.202 (0.168, 0.236)*** |
| Pre-pandemic slope | -0.002 (-0.007, 0.003) | -0.006 (-0.015, 0.003) |
| Post-emergence intercept change | -0.149 (-0.196, -0.101)*** | 0.135 (0.060, 0.210)*** |
| Post-emergence slope change | 0.037 (0.004, 0.070)* | -0.086 (-0.133, -0.038)*** |
| **Proportion of Inpatients Admitted to the ICU or NICU** | | |
| Pre-pandemic intercept | 0.263 (0.245, 0.282)*** | 0.260 (0.233, 0.288)*** |
| Pre-pandemic slope | 0.021 (0.017, 0.026)*** | -0.001 (-0.008, 0.006) |
| Post-emergence intercept change | -0.160 (-0.199, -0.121)*** | -0.038 (-0.098, 0.022) |
| Post-emergence slope change | -0.006 (-0.031, 0.019) | 0.010 (-0.028, 0.048) |
| **Proportion of Inpatients Mechanically Ventilated** | | |
| Pre-pandemic intercept | 0.120 (0.104, 0.135)*** | 0.117 (0.096, 0.138)*** |
| Pre-pandemic slope | 0.003 (-0.001, 0.007) | 0.001 (-0.004, 0.006) |
| Post-emergence intercept change | -0.103 (-0.141, -0.065)*** | -0.037 (-0.081, 0.008) |
| Post-emergence slope change | 0.030 (0.004, 0.056)* | -0.010 (-0.038, 0.018) |

Supplementary Table 3

**Supplementary Table 3.** Percent decline in the proportion of patients experiencing each outcome, by age group, from the pre-pandemic to the post-emergence phase. Age- and phase-stratified proportions are depicted in **Figure 4**.

| **Metric** | **Age Group** | **Percent Decline** |
|---|---|---|
| ED Admission | 0-3 months | 7.61 |
| ED Admission | 3-6 months | 8.36 |
| ED Admission | 6-12 months | 16.43 |
| ED Admission | 1-2 years | 25.61 |
| ED Admission | 2-4 years | 38.50 |
| ED Admission | 5-17 years | 46.08 |
| ICU or NICU Admission | 0-3 months | 2.86 |
| ICU or NICU Admission | 3-6 months | 11.39 |
| ICU or NICU Admission | 6-12 months | 16.39 |
| ICU or NICU Admission | 1-2 years | 19.54 |
| ICU or NICU Admission | 2-4 years | 22.96 |
| ICU or NICU Admission | 5-17 years | 25.23 |
| Mechanical Ventilation | 0-3 months | 17.03 |
| Mechanical Ventilation | 3-6 months | 39.96 |
| Mechanical Ventilation | 6-12 months | 43.01 |
| Mechanical Ventilation | 1-2 years | 46.20 |
| Mechanical Ventilation | 2-4 years | 52.93 |
| Mechanical Ventilation | 5-17 years | 46.68 |

#### Supplementary Figures

##### Supplementary Figure 1

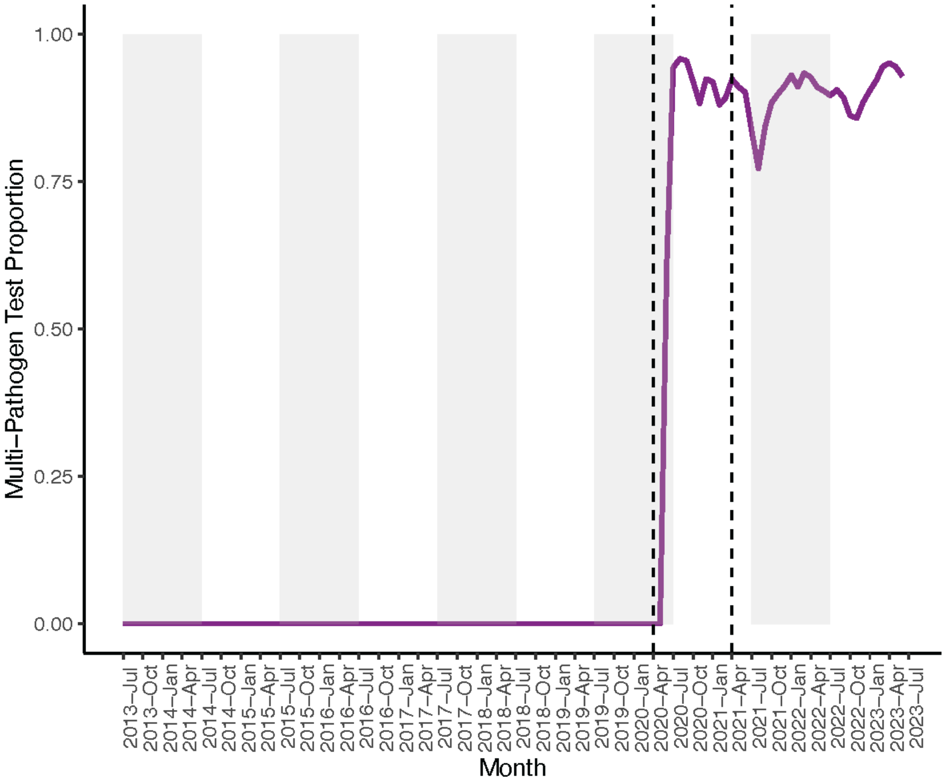


**Supplementary Figure 1.** Proportion of monthly respiratory syncytial virus (RSV) tests that were multi-pathogen tests. Single-pathogen tests are specific for RSV, while multi-pathogen tests are combination respiratory virus tests that also test for SARS-CoV-2. Gray rectangles denote alternating years (from July – July). Dashed lines delineate the end of the pre-pandemic phase and the start of the post-emergence phase.

Supplementary Figure 2

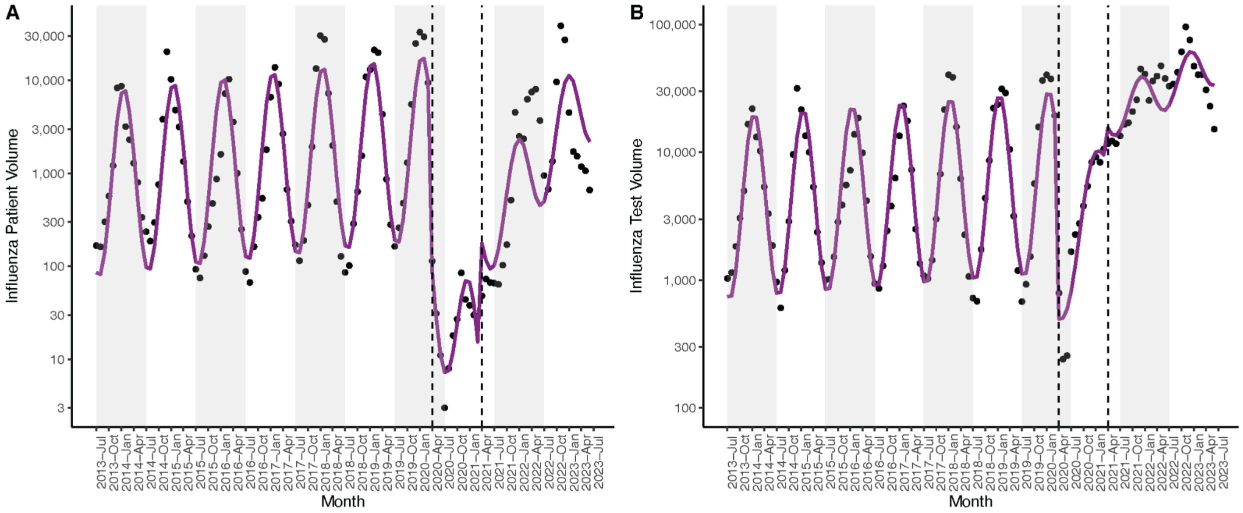


**Supplementary Figure 2.** Monthly influenza patient volume (**A**) and test volume (**B**). Black dots signify observed values, with model fit in magenta. Gray rectangles denote alternating years (from July – July). Dashed lines delineate the end of the pre-pandemic phase and the start of the post-emergence phase.

Supplementary Figure 3

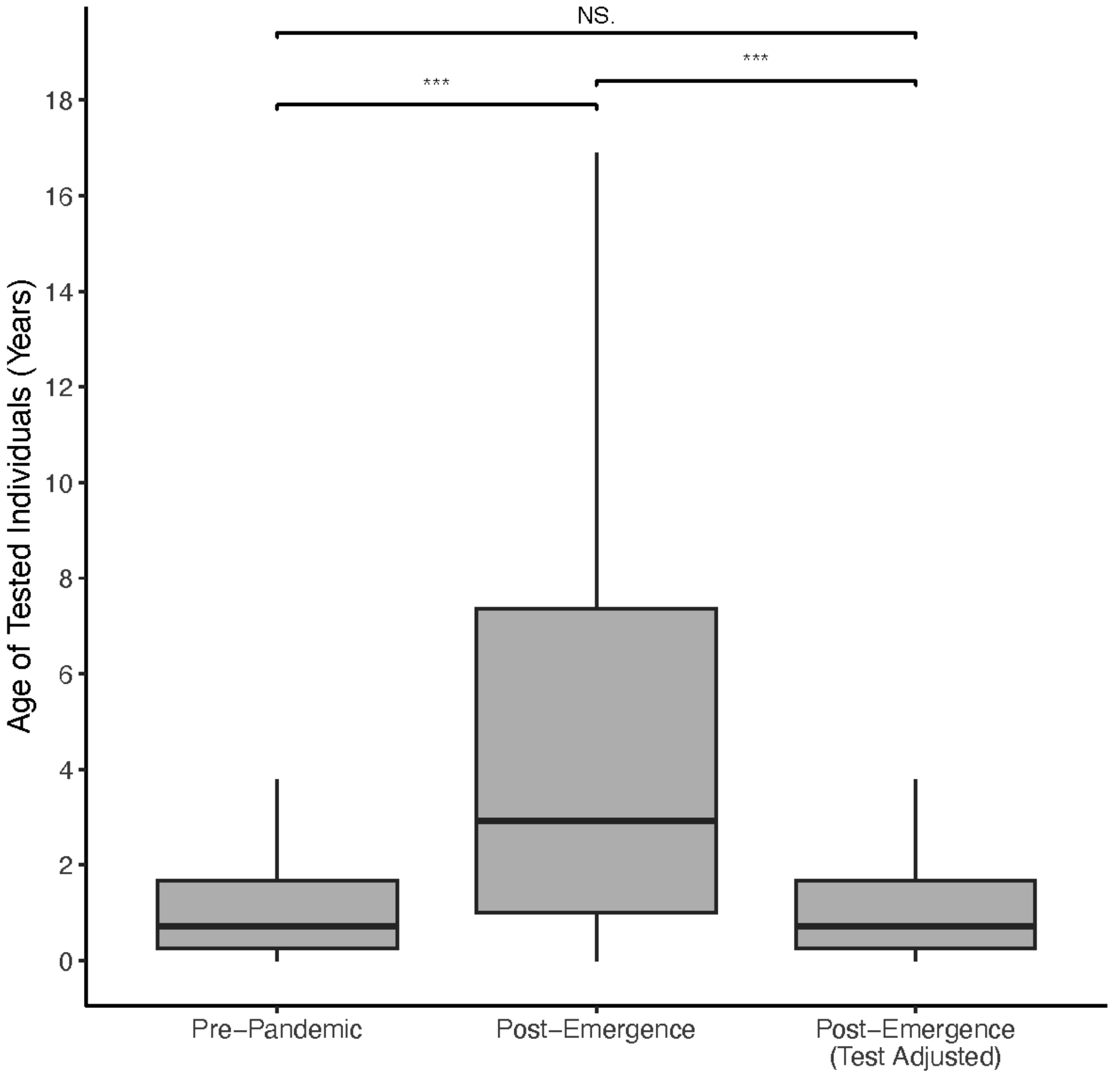


**Supplementary Figure 3.** Age, in years, of patients tested for RSV in the pre-pandemic and post-emergence phases. Because tests in the post-emergence phase were conducted on older patients, on average, than in the pre-pandemic phase, the post-emergence testing data was bootstrapped according to the age distribution of the pre-pandemic testing data. The resulting age distribution is reported as “Post-Emergence (Test Adjusted).” Boxplots display the first, second, and third quartiles, with whiskers extending to the minimum of 1.5 times the interquartile range and the most extreme data point in either direction. P-values via Wilcoxon rank sum test. ***, $p < 0.001$. NS, not significant.

Supplementary Figure 4

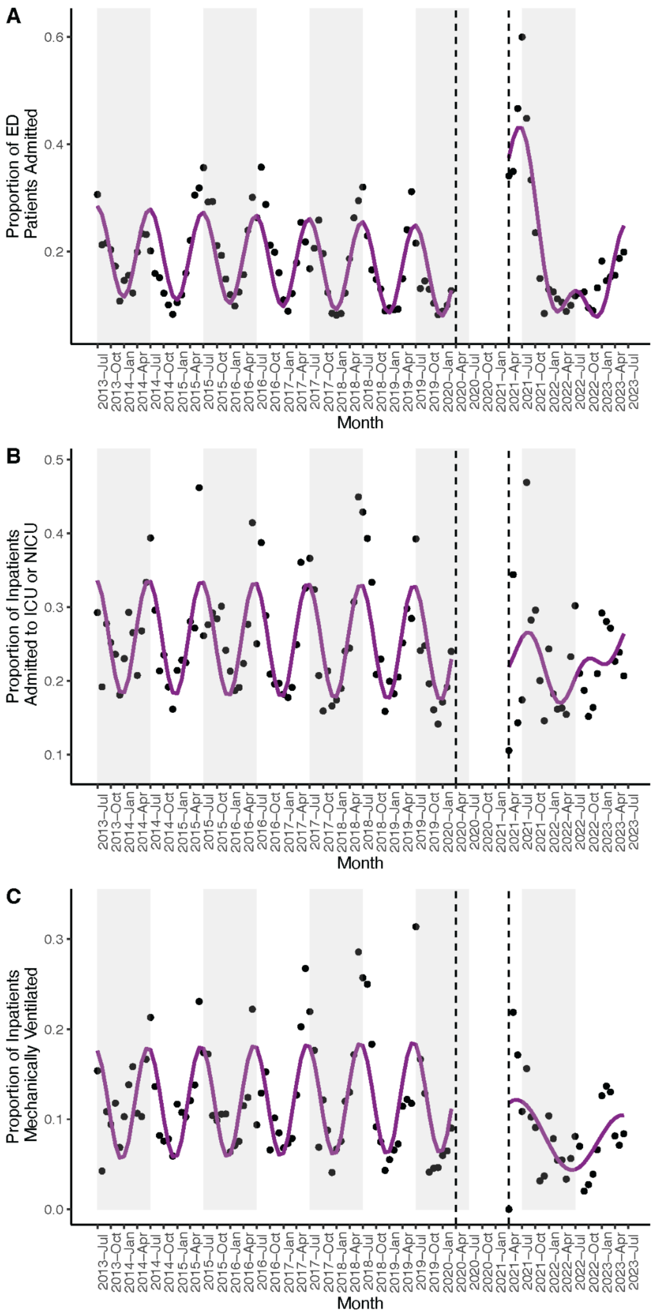


**Supplementary Figure 4. A.** Proportion of patients admitted from the emergency department (ED) for encounters with an influenza diagnosis. **B.** Proportion of inpatients with an influenza diagnosis admitted to the intensive care unit (ICU) or neonatal intensive care unit (NICU) **C.** Proportion of inpatients with an influenza diagnosis mechanically ventilated. **A-C.** Black dots signify observed values, with model fits in magenta. Gray rectangles denote alternating years (from July – July). Dashed lines delineate the end of the pre-pandemic phase and the start of the post-emergence phase. Data in the interim period is not shown.
